# Early Hemoglobin kinetics in response to ribavirin: Safety lesson learned from Hepatitis C to CoVID-19 therapy

**DOI:** 10.1101/2020.06.29.20142281

**Authors:** Antonio Rivero-Juarez, Mario Frias, Isabel Machuca, Marina Gallo, Pedro Lopez-Lopez, Angela Camacho, Antonio Rivero

## Abstract

**Background:** Ribavirin (RBV) is been used for SARS-CoV-2 infection. This drug is associated with a wide range of side effects, mainly anemia, so its use in patients with potential respiratory affectation could not be appropriate. The evidences of adverse events associated with RBV-use has mainly been derived in the context of hepatitis C (HCV) treatment, however the possible use of RBV in CoVID-19 patients could be limited to 14 days.

**Methods:** Longitudinal study including HIV/HCV coinfected patients. We evaluate the hemoglobin dynamics and reductions as well as evaluate the development rate of anemia during the first 2 weeks of therapy in HCV infected patients.

**Results:** 189 patients were included in the study. The median hemoglobin levels were 14.6 g/dL (IQR: 13.2-15.6 g/dL) and 13.5 g/dL (IQR: 12.3-14.5 g/dL) at weeks 1 and 2 of therapy, respectively. A cumulative number of 27 (14.2%) patients developed anemia (23 grade 1 [12.1%] and 4 grade 2 [2.1%]). We identify a baseline hemoglobin levels of 14 g/dL as the better cut-off to identify those patients with a high chance to develop anemia. Of the 132 patients with baseline hemoglobin level >14 g/dL, 8 developed anemia (6.1%) compared with 19 of 57 (33.3%) with hemoglobin levels lower than 14 g/dL (p < 0.001).

**Conclusions:** Our study shows valuable information about the early hemoglobin kinetic timing in patients on RBV-therapy, that could be useful to tailor CoVID-19 treatment if RBV use is considered.

## 1. Introduction

The management of the novel coronavirus (SARS-CoV-2) is challenging. The lack of direct acting drugs together with the absence of randomized clinical trials demonstrating the clinical benefit of repositioned drugs use, seriously compromised the virological control [1]. For this reason, the registration of clinical trials evaluating different drug combinations is increasing. The early expected results are those regarding the use of well-known drugs in combination with other compounds due to their demonstrated efficacy in reducing viral replication *in vitro* [2, 3]. One of these drugs is ribavirin (RBV), a guanosine analog that interferes with the RNA and DNA synthesis of several viruses. Due to its antiviral activity and the lack of specificity, RBV has been/is used for the treatment of several viral disease, such as hepatitis C (HCV), hepatitis E, or Crimea-Congo hemorrhagic fever [4-6]. Furthermore, RBV alone or in combination with other drugs has been used for the treatment of coronavirus such as SARS-CoV and MERS-CoV [7-10], including the SARS-CoV-2 [11].

RBV use is associated with a wide range of side effects, mainly hemolysis and the development of anemia [12]. Consequently, its use in patients with potential respiratory affectation and hypoxia could not be appropriate. The evidences of adverse events associated with RBV-use has mainly been derived in the context of HCV treatment, which could be as longer as 48 weeks. Because the possible use of RBV in CoVID-19 patients could be limited to 14 days, according to registered clinical trials [11], hemolysis and the rate of anemia development could be lower than previously reported due to a shorter therapy course. For this reason, the objective of our study was to evaluate the hemoglobin dynamics and reductions as well as evaluate the development rate of anemia during the first 2 weeks of therapy in HCV infected patients.

## 2. Materials and methods

### 2.1. Population

We retrospectively analyzed HIV/HCV patients prospectively included in the HCV therapy cohort of the Hospital Universitario Reina Sofía de Córdoba (Spain) that was established in January 2006 and February 2013. All patients started therapy treated with Peg-IFN α2a at doses of 180 μg, combined with a weight-adjusted dose of oral RBV (1000 mg/day for <75 kg, 1200 mg/day for ≥75 kg). Cirrhotic patients with a Child-Pugh Score of B or C were excluded.

### 2.2. Hemoglobin evaluation

All patients underwent hematological evaluation at each visit, including at baseline and at weeks 1, 2, 4, and every 4 weeks up to end of therapy. For the proposed of the study we only included hemoglobin values at baseline and at weeks 1 and 2. Hemoglobin decline was calculated from baseline to weeks 1 and 2. Normal hemoglobin levels were considered as hemoglobin levels between 18 and 12 g/dL. Only patients with normal hemoglobin levels at baseline (higher than 12 g/dL) were included in the study. Anemia was graded as, grade 1 (hemoglobin value between 11.9 and 10 g/dL), grade 2 (hemoglobin value between 9.9-8 g/dL), grade 3 (hemoglobin value between 7.9 and 6.5 g/dL), and grade 4 (hemoglobin value lower than 6.5 g/dL).

### 2.3. Statistical Analysis

The endpoint variable of the study was the development of anemia. Continuous variables were expressed as means ± standard deviation or median and quartiles (Q1-Q3) and were compared using Students t test or Mann-Whitney u test. Categorical variables were expressed as number of cases (percentages) and compared by Fisher exact test of Chi-square test. A multivariate logistic analysis was employed to identify independent variables associated with main outcome variable. Data were analyzed using the SPSS statistical software package release 18.0 (IBM Corporation, Somers, NY, USA and GraphPad Prism version 6 (Mac OS X version; GraphPad Software; San Diego, California, USA).

### 2.4. Ethical aspects

The study was designed and performed according to the Declaration of Helsinki. All patients provided written informed consent before included in the cohort. The study was approved by the Comité de Ética de Investigación de Córdoba (D439/2010). All patients signed an Informed consent form in writing that gave permission to store and process their samples in the Biobanco del Hospital Universitario Reina Sofía de Córdoba (ISCIII reference: B.0000419), which is integrated into the Biobanco del Sistema Sanitario Público de Andalucía, as well as to use of their clinical data for research.

## 3. Results

### 3.1. Population

A total of 189 patients were included in the analysis. Of them 150 (79.4%) were male, and the median age of was 43.3 years (IQR: 40.5-47.1 years). Fifty-one (27%) patients presented liver cirrhosis. The number of patients with available data at weeks 1 and 2 were 160 (84.6%) and 178 (94.2%), respectively.

### 3.2. Hemoglobin levels and reduction during the first two weeks of therapy

The median hemoglobin level at baseline was 15 g/dL (IQR: 13.9-15.7 g/dL). No differences were found in baseline hemoglobin levels between cirrhotic and noncirrhotic patients (14.5 g/dL [SD: 1.4 g/dL] vs. 15 g/dL [1.2 g/dL]; p = 0.083). The median hemoglobin levels were 14.6 g/dL (IQR: 13.2-15.6 g/dL) and 13.5 g/dL (IQR: 12.3-14.5 g/dL) at weeks 1 and 2 of therapy, respectively (Figure 1A). This indicated median hemoglobin kinetics of −0.1 g/dL (IQR: 0.3; −0.7 g/dL) at week 1, and 1.3 (IQR: − 0.5; −2.1 g/dL) at week 2 after the initiation of therapy (Figure 1B). No differences were observed between cirrhotic and noncirrhotic patients on mean hemoglobin reduction at week 1 (0.2 g/dL [IQR: 0.1-0.6 g/dL] vs. 0.1 g/dL [IQR: 0.3-0.8 g/dL]; p = 0.494) or week 2 (1.1 g/dL [IQR: 0.6-1.8 g/dL] vs. 1.3 g/dL [IQR: 0.4-2.3 g/dL]; p = 0.165). The RBV dose was not reduced in any of the patients during the study period. In Table 1, we show the number of patients with no hemoglobin reduction, reduction lower than 1 g/dL, reduction between 1 g/dL and 2 g/dL, and those showing a reduction of more than 2 g/dL at weeks 1 and 2 of therapy. At week 1 29 (18.1%) patients showed a hemoglobin reduction greater than 1 g/dL, while at week 2, 107 (60.4%) patients showed a hemoglobin reduction greater than 1 g/dL.

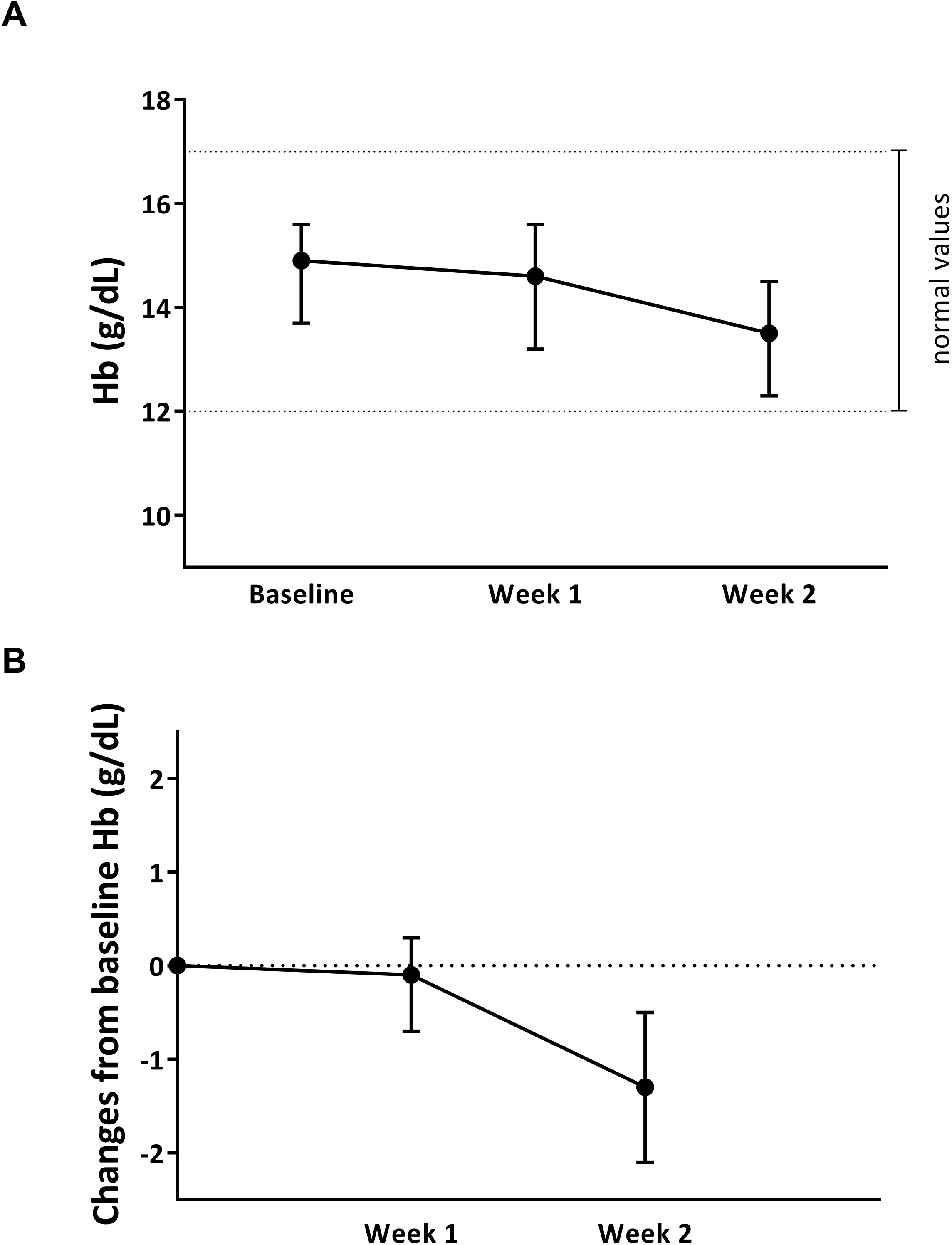

**Table 1.** Hemoglobin reduction during two weeks of ribavirin therapy.

|  | **Week 1 (n =160)** | **Week 2 (N = 178)** |
| --- | --- | --- |
| *No Hemoglobin reduction* | 70 (43.8%) | 22 (12.4%) |
| *Hemoglobin reduction < 1g/dL* | 61 (38.1%) | 49 (27.5%) |
| *Hemoglobin reduction 1-2 g/dL* | 25 (15.6%) | 55 (30.9%) |
| *Hemoglobin reduction >2 g/dL* | 4 (2.5%) | 52 (29.2%) |

### 3.3. Development of anemia

A total of 5 (2.6%) patients developed grade 1 anemia after 1 week of therapy. Among these patients, the median hemoglobin levels were 11.9 g/dL (IQR: 11-11.9 g/dL) at this point. At week 2, 18 (10.1%) and 4 (2.2%) patients showed grade 1 and 2 anemia, respectively. At this point, the median hemoglobin levels were 11.1 g/dL (IQR: 10.5-11.6 g/dL) and 9.75 g/dL (IQR: 9.7-9.8 g/dL) in patients with grade 1 and 2 anemia, respectively. Consequently, during the first two weeks of RBV therapy, a cumulative number of 27 (14.2%) patients developed anemia (23 grade 1 [12.1%] and 4 grade 2 [2.1%]). There was no reported treatment withdrawal during the study period due to adverse events.

### 3.4. Baseline factors associated with development of anemia

The median baseline hemoglobin levels were lower among patients who develop anemia during the treatment period than in those who did not (13.8 g/dL [IQR: 14.7-12.8 g/dL] vs. 15.1 g/dL [IQR: 15.9-14.1 g/dL]; p < 0.001). We identify a baseline hemoglobin levels of 14 g/dL as the better cut-off to identify those patients with a high chance to develop anemia. Of the 132 patients with baseline hemoglobin level higher than 14 g/dL, 8 develop anemia (6.1%) compared with 19 of 57 (33.3%) with hemoglobin levels lower or equal to 14 g/dL (p < 0.001). In Table 2 we shown the proportion of patients who develop grade 1 and 2 anemia according to baseline hemoglobin levels.

**Table 2.** Patients developing anemia, grade 1 anemia, and grade 2 anemia according to baseline hemoglobin levels

| Baseline Hemoglobin | N | Overall anemia | Grade 1 anemia | Grade 2 anemia |
| --- | --- | --- | --- | --- |
| *> 14 g/dL* | 132 | 8 (6.1%) | 7 (5.3%) | 1 (0.7%) |
| ***≤14 g/dL*** | 57 | 19 (33.1%) | 16 (28.1%) | 3 (5.2%) |
| *P value* |  | < 0.001 | < 0.001 | 0.082 |

By multivariate analysis, we identify a baseline hemoglobin level ≤ 14 g/dL as the main factor associated with development of anemia, adjusted by gender, age and liver cirrhosis (Table 3).

**Table 3.** Logistic multivariate analysis for the development of anemia

| **Variable** | **Condition** | **OR** | **95% CI** |
| --- | --- | --- | --- |
| **Age** |  | 0.989 | 0.913-1.013 |
| **Gender** | *Male* | 1 |  |
|  | *Female* | 0.754 | 0.278-2.049 |
| **Cirrhosis** | *No* | 1 |  |
|  | *Yes* | 0.954 | 0.357-2.546 |
| **Baseline Hemoglobin** | *> 14 g/dL* | 1 |  |
|  | *≤ 14 g/dL* | 7.491 | 2.789-20.123 |

## 4. Discussion

Because the lack of direct acting antiviral drugs for SARS-CoV-2, several drugs with demonstrated *in vitro* antiviral activity have been repositioned and are currently been evaluated in clinical trials [1, 2]. In this sense, RBV, due to its wide antiviral activity, is one of them. However, due to the high rate of adverse events related to the use of RBV, overall hemolysis, its use for the treatment of SARS-CoV-2 infection could be limited. Our study, using patients treated for HCV infection information, report the hemoglobin kinetic of two weeks of therapy, identifying a subgroup of patients with low likelihood to develop anemia.

Recently, the RBV efficacy to inhibit viral replication of SARS-CoV-2 has been evaluated *in vitro*, finding that the proper dose employed to achieve EC_50_ should not be lower than 100 uM [3]. Nevertheless, as authors noted, the lack of nucleoside kinases of the cell line employed in the study fail to convert the drug in to its triphosphate form, which have the main antiviral activity [13], so the dose required could be lower. In fact, RBV, alone or in combination with other compounds, at lower concentration that suggested *in vitro*, have shown to improve the clinical outcome of patients infected with SARS-CoV [7-9], and MERS-CoV [10]. However, because these are not randomized controlled trials evidences regarding efficacy cannot be considered. In contrast, lessons regarding to adverse events using RBV can be useful. In a Canadian study, 126 patients diagnosed of SARS-CoV received RBV within the first 48 hours after admission during a median time of 6 days (IQR: 5-7 days) using an initial dose of 2 g intravenously, followed by 1 g intravenously every 6 hours for 4 days, followed by 500 mg every 8 hours for 3 days [14]. In this study RBV was associated with significant toxicity, in sense that 49% of patients experienced a decrease in hemoglobin level of at least 2 g/dL and led to the premature discontinuation of RBV in 18% of patients. In the same way, another Canadian report including 110 patients, observed a significant decrease in the hemoglobin level after 6.8 days (SD =2.7 days) of RBV initiation at the same dose that the previous study, achieving the lowest hemoglobin level at day 13 (SD = 4.1 days) [15]. In these studies, the development of anemia was strongly associated with the use of high dose of RBV (i.e., a cumulative dose higher than 20 g), supposing 40 of 59 patients (67.7%). Although obvious differences between populations compared, the patients analyzed in our study showed a similar timing of hemoglobin kinetic after starting therapy, cumulating at the second week of therapy the appearance of anemia. In our study, the percentage of patients with anemia compared with this study could be related to the lower dose employed, which is in fact a similar dose used in clinical trials for SARS-CoV-2 (clinicaltrial.gov identifier: NCT04276688).

Our study shows valuable information about the early hemoglobin kinetic timing in patients on RBV-therapy, that could be useful to tailor CoVID-19 treatment if RBV use is considered. We identify a subgroup of patients with a low likelihood to develop anemia during 14 days of therapy with RBV. In this sense, those patients showing a baseline hemoglobin level higher than 14 g/dL could be considered as low-risk population, with an anemia rate of 6.1%. Our results suggest that, in this subset of patients with RBV could be use with relatively safety.

## Data Availability

Raw data is available under reasonable request to corresponding author

## References

[1] Totura AL, Bavari S. Broad-spectrum coronavirus antiviral drug discovery. Expert Opin Drug Discov 2019; 14:397–412.

[2] Mungroo MR, Khan NA, Siddiqui R. Novel Coronavirus: Current Understanding of Clinical Features, Diagnosis, Pathogenesis, and Treatment Options. Pathogens 2020; 9: E297.

[3] Choy KT, Wong AY, Kaewpreedee P, et al. Remdesivir, lopinavir, emetine, and homoharringtonine inhibit SARS-CoV-2 replication in vitro. Antiviral Res 2020; 178: 104786.

[4] Feld JJ, Jacobson IM, Sulkowski MS, Poordad F, Tatsch F, Pawlotsky JM. Ribavirin revisited in the era of direct-acting antiviral therapy for hepatitis C virus infection. Liver Int 2017; 37: 5–18.

[5] de la Calle-Prieto F, Martín-Quirós A, Trigo E, et al. Therapeutic management of Crimean-Congo haemorrhagic fever. Manejo terapéutico de la fiebre hemorrágica de Crimea-Congo. Enferm Infecc Microbiol Clin 2018; 36: 517–522.

[6] Rivero-Juarez A, Vallejo N, Lopez-Lopez P, et al. Ribavirin as a First Treatment Approach for Hepatitis E Virus Infection in Transplant Recipient Patients. Microorganisms 2019; 8: 51.

[7] Peiris JS, Chu CM, Cheng VC, et al. Clinical progression and viral load in a community outbreak of coronavirus-associated SARS pneumonia: a prospective study. Lancet 2003; 361: 1767–1772.

[8] Lee N, Hui D, Wu A, et al. A major outbreak of severe acute respiratory syndrome in Hong Kong. N Engl J Med 2003; 348: 1986–1994.

[9] Poutanen SM, Low DE, Henry B, et al. Identification of severe acute respiratory syndrome in Canada. N Engl J Med 2003; 348: 1995–2005.

[10] Arabi YM, Al-Omari A, Mandourah Y, et al. Critically Ill Patients With the Middle East Respiratory Syndrome: A Multicenter Retrospective Cohort Study. Crit Care Med 2017; 45: 1683–1695.

[11] Khalili JS, Zhu H, Mak NSA, Yan Y, Zhu Y. Novel coronavirus treatment with ribavirin: Groundwork for an evaluation concerning COVID-19. J Med Virol 2020; 10.1002/jmv.25798. doi:10.1002/jmv.25798

[12] Brennan BJ, Wang K, Blotner S, et al. Safety, tolerability, and pharmacokinetics of ribavirin in hepatitis C virus-infected patients with various degrees of renal impairment. Antimicrob Agents Chemother 2013; 57: 6097–6105.

[13] Thomas E, Ghany MG, Liang TJ. The application and mechanism of action of ribavirin in therapy of hepatitis C. Antivir Chem Chemother 2012; 23: 1–12.

[14] Booth CM, Matukas LM, Tomlinson GA, et al. Clinical features and short-term outcomes of 144 patients with SARS in the greater Toronto area. JAMA 2003; 289: 2801–2809.

[15] Knowles SR, Phillips EJ, Dresser L, Matukas L. Common adverse events associated with the use of ribavirin for severe acute respiratory syndrome in Canada. Clin Infect Dis 2003; 37: 1139–1142.

